# Deteriorated mechanics of left ventricular diastolic filling one year after coronary artery bypass grafting

**DOI:** 10.1101/2024.11.28.24318125

**Authors:** Joakim Norderfeldt, Martin G Sundqvist, Eva Maret, Ulrika Löfström, Matthias Corbascio, Camilla Hage, Mattias Ekström, Håkan Wallén, Patrik Lyngå, Bengt Persson, Hans E Persson, Cecilia Linde, David Marlevi, Maria J Eriksson, Martin Ugander

**Author notes:** Corresponding author: Address for correspondence, Martin Ugander, Kolling Building, Level 12 Royal North Shore Hospital, St Leonards, Sydney, NSW 2065 Australia. The study was performed at Karolinska Institutet, Stockholm, Sweden. Shared last author (David Marlevi, Maria J Eriksson, Martin Ugander).

## Abstract

**Background:** Ischemic heart disease impairs left ventricular (LV) diastolic function, but little is known about changes in the mechanical properties of LV relaxation following coronary artery bypass grafting (CABG).

**Objectives:** This study aimed to explore if and how the mechanics of LV filling change following CABG

**Methods:** Patients underwent transthoracic echocardiography before and one year after elective CABG. Mitral inflow Doppler E-waves were analysed using the parameterized diastolic filling (PDF) method, allowing for quantification of diastolic filling mechanics.

**Results:** Among patients (n=96, 10% female, median [interquartile range] age 68 [62–74] years), LV ejection fraction (LVEF) at baseline was 59 [53–63] %. At follow-up, there was an increase in the PDF-derived measures of myocardial stiffness, damping, peak driving and resistive forces, together with increase in left atrial (LA) volume index, and a decrease in LA conduit and contractile strains and tricuspid annular plane systolic excursion (p<0.001 for all). There was no change in heart rate, LV size, LVEF, E/é ratio, or LV filling efficiency. Furthermore, there were no differences in the mechanics of LV filling in patients when grouped according to baseline LVEF, or the number of revascularized coronary arteries

**Conclusions:** One year after CABG, there was consistent deterioration in the mechanics of diastolic filling assessed using mechanistic evaluation. Revascularization with CABG does not improve diastolic function one year after CABG. However, causes and significance of these changes remain to be further investigated. The PREFERS study is registered at clinicaltrials.gov with ID-number NCT03671122.

**Graphical abstract:** 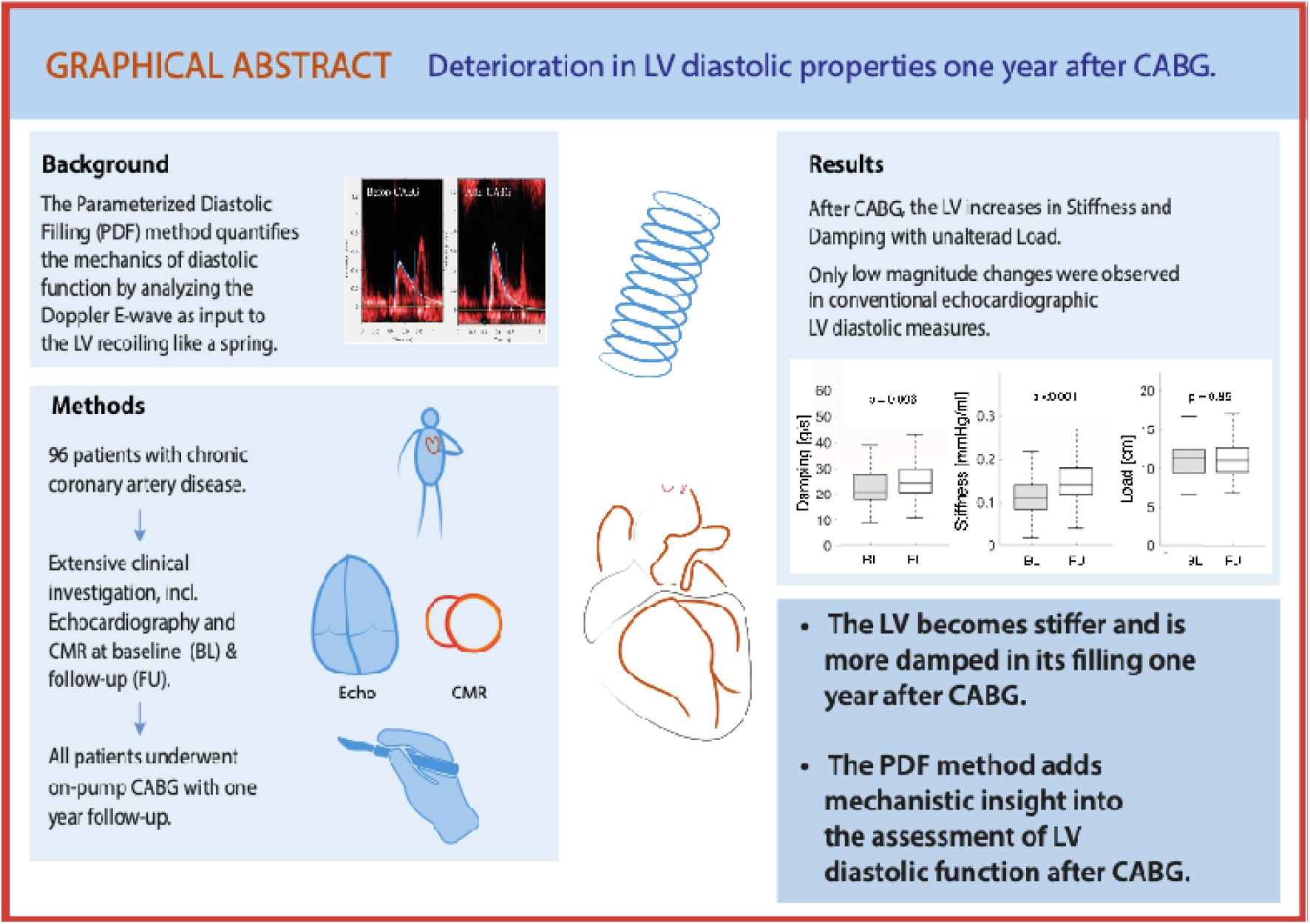

Overview of background, study design, methods and main results.

## 1. Introduction

Ischemic heart disease gives rise to a broad range of symptoms and myocardial changes driven by different pathophysiological processes. Diastolic dysfunction is one of the first manifestations of impaired cardiac function due to myocardial ischemia^1^.

Left ventricular (LV) diastolic function is characterized by LV relaxation, stiffness, and early diastolic recoil. All of these factors influence LV filling pressure. Furthermore, LV diastolic dysfunction has been shown to impact both patient symptoms and outcomes^2^. Echocardiography is a cornerstone in the evaluation of LV diastolic properties and function^2^.

Current clinical guidelines on echocardiographic evaluation of LV diastolic function^3^ aim to determine whether LV filling pressure is elevated or not. However, such analysis does not consider the inherent mechanical characteristics of the myocardium, with elevated myocardial stiffness a quantifiable marker of progressive heart failure^4, 5^. The echocardiographic parameterized diastolic filling (PDF) method is a validated approach for assessing the mechanics of LV diastolic filling. The method uses the velocity profile of early diastolic transmitral flow (the E-wave) from pulsed-wave Doppler echocardiography to quantify mechanical stiffness, damping, and load^6, 7^, together providing quantitative information on intrinsic mechanical properties of the LV. Stiffness contributes to the forces *driving* the recoil of the LV, whereas damping contributes to the forces *resisting* the recoil of the LV. Load is related to the stroke volume, and is closely correlated with the E-wave velocity time integral^7^ (see **Table 1**). The PDF method has up until now been used to illustrate mechanical differences in cardiac relaxation in clinical situations such as patients with diabetes mellitus and hypertension^10, 15, 16^.

**Table 1.** PDF parameters with explanation of physiological basis and clinical relevance.

| Name | Parameter | Unit (SI) | Physiological basis and clinical relevance |
| --- | --- | --- | --- |
| Stiffness | $k$ | g/s<sup>2</sup> (N/m)<br>mmHg/ml | The force per unit displacement with which the LV restores its volume during early diastolic filling<br>Contributes to the restoring force that *drives* early filling (Peak driving force of filling, $kx_o$)<br>Linearly related to invasively measured chamber stiffness in mmHg/ml ($dP/dV$)<sup>8</sup><br>Increased in diabetes and commonly also in both constrictive and restrictive filling<sup>9</sup> |
| Damping | $c$ | g/s (N·s/m) | The viscous energy loss that counteracts LV recoil during early diastolic filling<br>Contributes to the resistive force that *opposes* early filling (Peak resistive force of filling, $cE$)<br>Reflects impaired relaxation and increased myocardial viscoelasticity<br>Non-linearly related to the time constant of LV isovolumic pressure decay ($Tau$, $\tau$)<br>Increased in diabetes<sup>9</sup>, hypertension<sup>9, 10</sup> and aortic stenosis<sup>11</sup> |
| Load | $x_{\square}$ | cm | The volume load that compresses the myocardium at end systole<br>Proportional to the E-wave velocity-time integral and the early component of the LV filling volume |
| E-wave peak velocity | $E$ | m/s | The peak velocity of blood flow across the mitral valve during early LV filling |
| Peak driving force of filling | $kx_{\square}$ | mN | The peak force driving LV filling<br>The product of stiffness ($k$) and load ($x_{\square}$)<br>Analogous to the peak atrioventricular pressure gradient<sup>12</sup> |
| Peak resistive force of filling | $cE$ | mN | The peak force opposing LV filling<br>The product of damping ($c$) and the E-wave peak velocity ($E$)<br>Represents the maximum viscoelastic resistive force during filling<sup>13</sup> |
| Time constant of isovolumic pressure decay | $\tau$ | ms | Time constant describing the exponential rate at which LV pressure declines before mitral valve opening<br>Can be estimated by a non-linear combination of damping ($c$) and stiffness ($k$)<br>Increased in impaired relaxation<sup>14</sup> |

2 *Legend:* Key PDF parameters with mechanistic background and clinical significance.

Beyond optimal medical therapy, patients diagnosed with ischemic heart disease benefit from surgical revascularization with coronary artery bypass graft (CABG) surgery^17^. Whilst CABG is known to alleviate regional stress-induced myocardial ischemia, by comparison, long-term changes in LV diastolic function after CABG have been scarcely studied^7, 18, 19^. During the first hour after CABG, it is known that there is an increase in LV stiffness and reduced LV compliance^18^, and the extent of deterioration in diastolic function has been associated with the duration of aortic cross clamp^18^. Furthermore, an immediate leftward shift in the curve of the LV end-diastolic pressure to LV end-diastolic diameter has been reported after CABG^18^. However, the intrinsic mechanical properties of LV diastolic function quantified by the PDF method have not yet been studied after CABG. Therefore, the aim of this study was to evaluate if and how the mechanical properties of LV diastolic function were affected one year after CABG using the PDF method.

## 2. Methods

### 2.1 Study population

The current study is a substudy of CABG PREFERS (Preserved and Reduced Ejection Fraction Epidemiological Regional Study) performed in Stockholm, Sweden, as previously described^20^. Briefly, all patients admitted for elective isolated CABG surgery with or without previous acute myocardial infarction (AMI) or history of heart failure at the Department of Cardiothoracic Surgery at Karolinska University Hospital, Stockholm, Sweden, between March 2016 and January 2020, were asked to participate. Inclusion and exclusion criteria have been described in detail in the original study design publication. At enrolment and one year after CABG, patients underwent comprehensive examination including ECG, blood sampling, and transthoracic echocardiography (TTE). The study complied with the Declaration of Helsinki and was approved by the regional ethics committee in Stockholm, Sweden (DNR 2013/1869-31/1). All patients provided written informed consent prior to participation.

Before surgery, the perioperative risk was assessed using the risk stratification score EuroSCORE 2. CABG operations (n=96) were performed with patients on cardiopulmonary bypass according to clinical routine. All but three were performed at the Karolinska University Hospital, Stockholm, with three performed at the Örebro University Hospital, Örebro. Ninety patients were operated on by the same consultant surgeon. Revascularization was accomplished using double mammary arterial grafting and, in some cases also by venous grafts, where applicable. The duration of cardiopulmonary bypass and aortic cross-clamp were collected from medical records.

### 2.3 Echocardiography

TTE image acquisition and analysis was performed according to clinical recommendations^21, 22^ by two experienced sonographers using Vivid E95 (General Electric, Horten, Norway). All echocardiographic examinations were analysed by two experienced specialists in cardiovascular imaging (MJE and EM) blinded to timepoint of examination, using commercial software (EchoPac, General Electric, Horten, Norway).

### 2.4 Parameterized diastolic filling (PDF)

PDF measurements quantitatively describe the intrinsic mechanical properties of the LV in analogy to that of a recoiling spring, and has previously been described^6, 7^. In brief, the analysis results in three main parameters; stiffness (*k*), damping (*c*) and load (*x_0_*). From these main parameters, additional parameters can be derived, all representing different aspects of the LV recoil, e.g. peak driving force and peak resistive force. For analysis of mitral E-waves by the PDF method, pulsed wave Doppler of the mitral inflow with a horizontal sweep speed of 100 mm/s was collected for 30 seconds during free breathing. If there was fusion of the E- and the A-wave, the examination was excluded if the fusion occurred at or above 50 % of the maximum E-wave velocity, as has previously been recommended^7^. All PDF analyses were performed using the freely available software Echo E-waves (www.echoewaves.org, version 1.01), see **Graphical abstract**. PDF analysis was performed blinded to knowledge of baseline or follow-up timepoint. PDF measures of LV myocardial stiffness are reported as mmHg/ml after unit conversion as previously described^8^.

### 2.6 Cardiac phenotype definitions

Patients were divided into phenotypes of Normal, preserved EF (pEF), or reduced EF (rEF) following pre-specified approach and criteria using echocardiographic findings and N-terminal pro b-type natriuretic peptide (NT-proBNP) concentrations as previously described^20^, in brief using the ratio of E/e’ together with left atrial volume index (LAVI) and in equivocal cases determined by consensus between two experts (MJE and HP).

### 2.7 Statistical analysis

Results are presented as median [interquartile range], with statistical significance set at p<0.05. Differences between baseline and follow-up were tested using the Wilcoxon signed rank test with continuity correction. Only patients with available data at both baseline and follow-up were included. If missing values, the number of patients included in the analysis is noted in respective tables. For subgroup analyses, comparison between groups were performed using the Kruskal-Wallis test. For differences between the phenotype groups regarding echocardiography and NT-proBNP, the Mann Whitney U-test was used. The change in mechanical properties (PDF) within each cardiac phenotype group was analysed using Wilcoxon signed rank sum test. Variable changes were correlated to the duration of cardiopulmonary bypass, aortic cross-clamp time, and preoperative Euroscore2 using the Spearman correlation coefficient. Statistical analyses were performed using SPSS (IBM SPSS Statistics, New York, USA), version 28.0.0.0.

## 3. Results

### 3.1 Patient population

138 patients were included in CABG PREFERS. Out of these, patients were excluded based on insufficient image quality to perform PDF analysis at either baseline or follow-up (n=15), absence of follow-up examination (n=15), absence of CABG surgery (n=9), admission for acute surgery (n=1), concomitant valve surgery (n=1) and transfer to another hospital for surgery (n=1).

Out of the 96 remaining patients, 86 (90 %) were males with an age of 68 [62–74] years. **Table 2** summarizes the population characteristics. The time between baseline TTE and surgery was 63 [29–104] days. The interval between surgery and follow-up TTE was 363 [351–372] days.

**Table 2.** Patient characteristics at baseline.

|  |  |
| --- | --- |
| Total number of included patients, n | 96 |
| Male sex, n (%) | 86 (90) |
| Age, years | 68 [62–74] |
| Height, m | 1.75 [1.71–1.81] |
| Weight, kg | 83 [73–92] |
| BSA, m<sup>2</sup> | 1.95 [1.85–2.10] |
| BMI, kg/m<sup>2</sup> | 25.6 [23.5–29.0] |
| Hypertension, n (%) | 39 (72) |
| Previous infarction by LGE CMR<sup>§</sup>, n/n (%) | 48/78 (62) |
| Diabetes mellitus type I, n (%) | 1 (1) |
| Diabetes mellitus type II, n (%) | 30 (31) |
| Diabetes mellitus type unknown, n (%) | 1 (1) |
| Sinus rhythm at echocardiographic examination, n (%) | 90 (94) |
| Systolic blood pressure, mmHg | 144 [128-152] |
| Diastolic blood pressure, mmHg | 79 [73-87] |
| Patients treated with: |  |
| - Beta blocker, n (%) | 56 (58) |
| - ACE-I, n (%) | 23 (24) |
| - ARB, n (%) | 23 (24) |
| - ACE-I and ARB, n (%) | 2 (2) |
| - ARNI, n (%) | 0 (0) |
| - MRA, n (%) | 3 (3) |
| - CCB, n (%) | 27 (28) |
| - Long-acting nitrate, n (%) | 36 (38) |
| - Loop or thiazide diuretic, n (%) | 4 (4) / 5 (5) |

### 3.2 Echocardiographic parameters

Conventional echocardiographic parameters are presented in **Table 3**. A total of 14 (15%) patients had a baseline left ventricular ejection fraction (LVEF) below ≤45 %. LV diastolic volume at follow-up was slightly reduced, while LV systolic volume did not change. There was no difference in LVEF, mitral annular plane systolic excursion or LV global longitudinal strain between baseline and follow-up. No difference could be observed in right ventricular size between baseline and follow-up, but tricuspid annular plane systolic excursion was lower at follow-up. Left atrial (LA) reservoir strain and conduit strain were decreased at follow-up compared to baseline, but no change could be seen in LA contraction strain. NT-proBNP did not differ between baseline and follow-up.

**Table 3.**
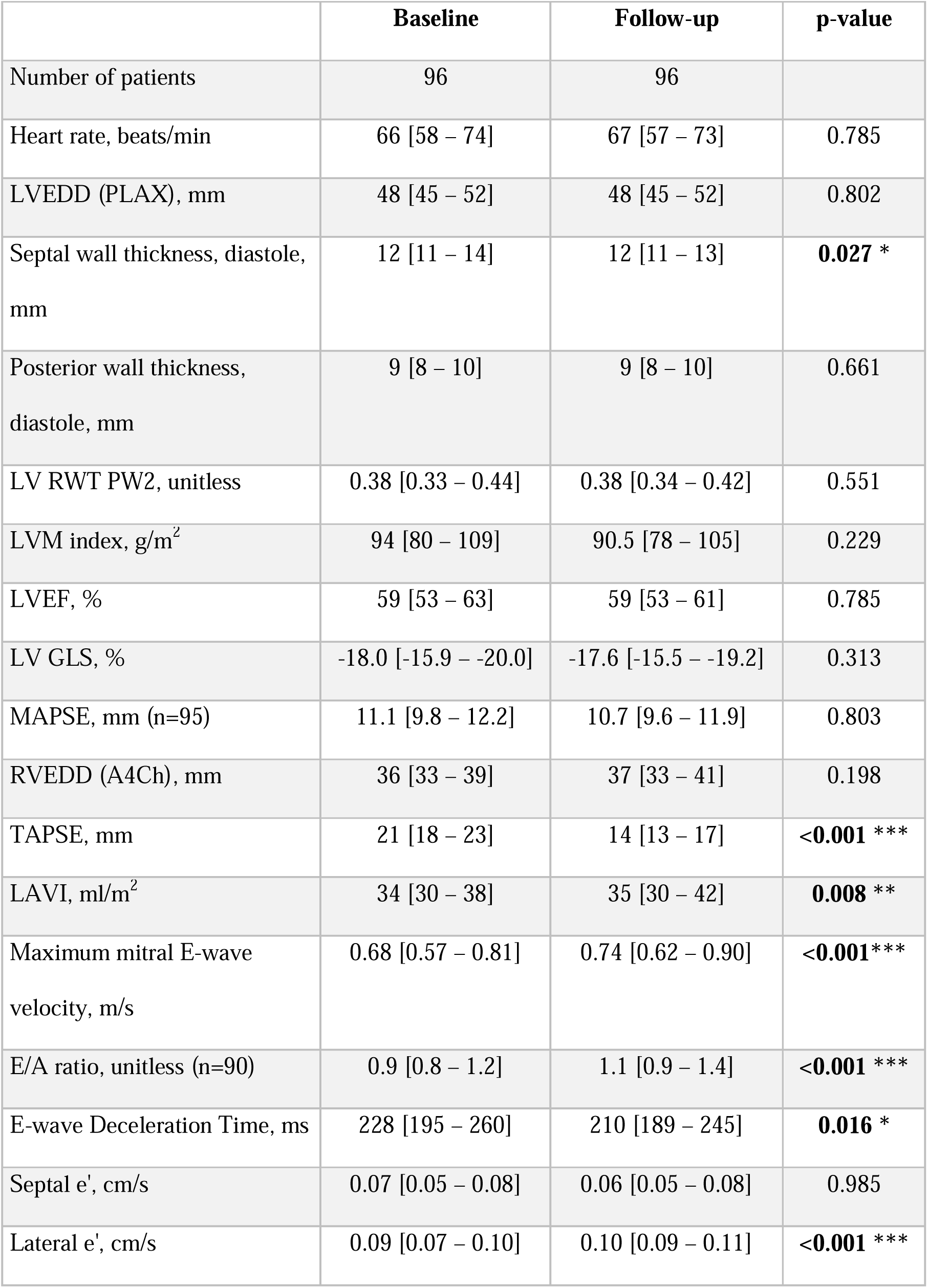
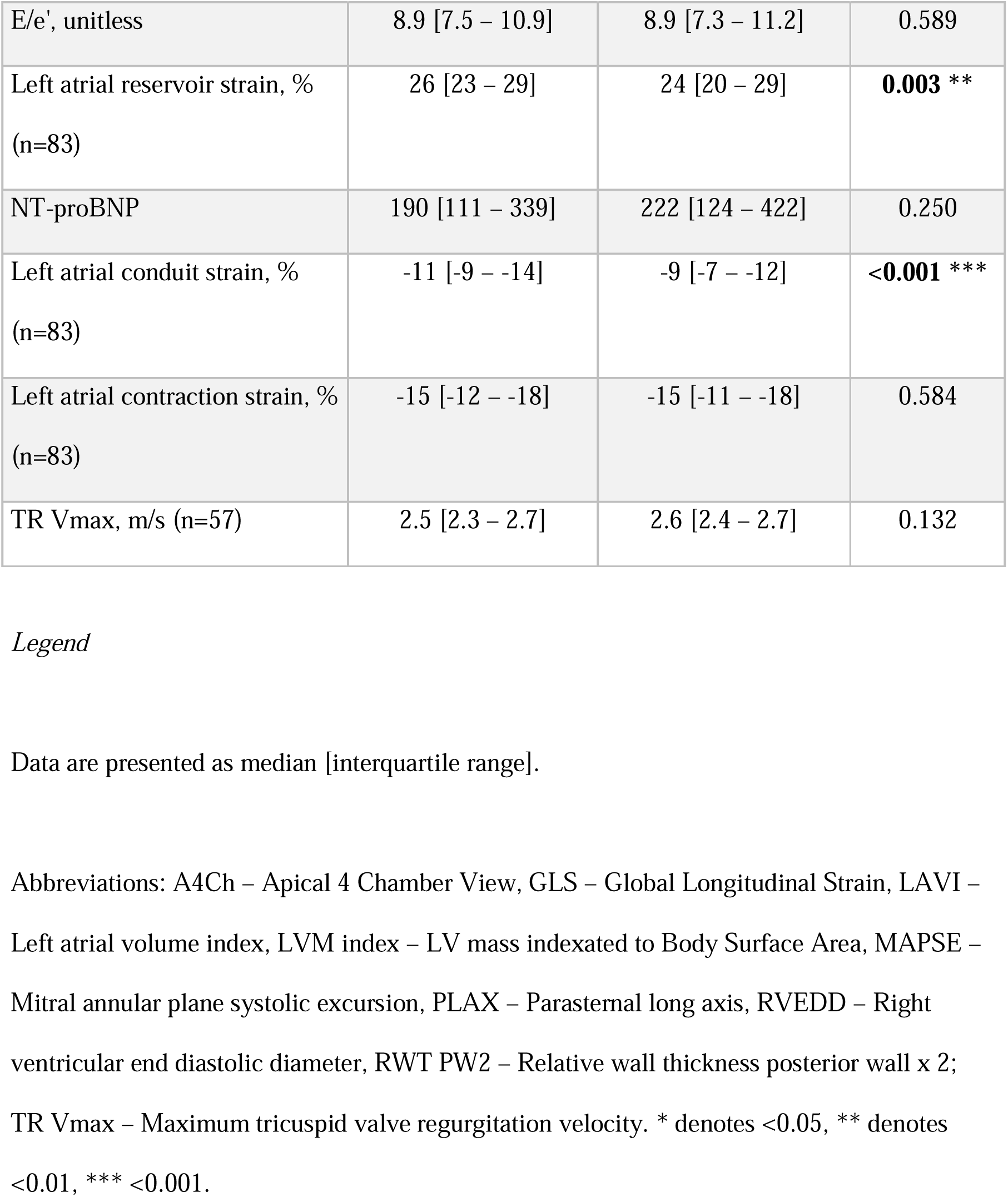
Conventional echocardiographic parameters at baseline and at follow-up one year after CABG.

|  | Baseline | Follow-up | p-value |
| --- | --- | --- | --- |
| Number of patients | 96 | 96 |  |
| Heart rate, beats/min | 66 [58 – 74] | 67 [57 – 73] | 0.785 |
| LVEDD (PLAX), mm | 48 [45 – 52] | 48 [45 – 52] | 0.802 |
| Septal wall thickness, diastole, mm | 12 [11 – 14] | 12 [11 – 13] | **0.027 \*** |
| Posterior wall thickness, diastole, mm | 9 [8 – 10] | 9 [8 – 10] | 0.661 |
| LV RWT PW2, unitless | 0.38 [0.33 – 0.44] | 0.38 [0.34 – 0.42] | 0.551 |
| LVM index, g/m<sup>2</sup> | 94 [80 – 109] | 90.5 [78 – 105] | 0.229 |
| LVEF, % | 59 [53 – 63] | 59 [53 – 61] | 0.785 |
| LV GLS, % | -18.0 [-15.9 – -20.0] | -17.6 [-15.5 – -19.2] | 0.313 |
| MAPSE, mm (n=95) | 11.1 [9.8 – 12.2] | 10.7 [9.6 – 11.9] | 0.803 |
| RVEDD (A4Ch), mm | 36 [33 – 39] | 37 [33 – 41] | 0.198 |
| TAPSE, mm | 21 [18 – 23] | 14 [13 – 17] | **<0.001 \*\*\*** |
| LAVI, ml/m<sup>2</sup> | 34 [30 – 38] | 35 [30 – 42] | **0.008 \*\*** |
| Maximum mitral E-wave velocity, m/s | 0.68 [0.57 – 0.81] | 0.74 [0.62 – 0.90] | **<0.001\*\*\*** |
| E/A ratio, unitless (n=90) | 0.9 [0.8 – 1.2] | 1.1 [0.9 – 1.4] | **<0.001 \*\*\*** |
| E-wave Deceleration Time, ms | 228 [195 – 260] | 210 [189 – 245] | **0.016 \*** |
| Septal e', cm/s | 0.07 [0.05 – 0.08] | 0.06 [0.05 – 0.08] | 0.985 |
| Lateral e', cm/s | 0.09 [0.07 – 0.10] | 0.10 [0.09 – 0.11] | **<0.001 \*\*\*** |
| E/e', unitless | 8.9 [7.5 – 10.9] | 8.9 [7.3 – 11.2] | 0.589 |
| Left atrial reservoir strain, %<br>(n=83) | 26 [23 – 29] | 24 [20 – 29] | **0.003 \*\*** |
| NT-proBNP | 190 [111 – 339] | 222 [124 – 422] | 0.250 |
| Left atrial conduit strain, %<br>(n=83) | -11 [-9 – -14] | -9 [-7 – -12] | **<0.001 \*\*\*** |
| Left atrial contraction strain, %<br>(n=83) | -15 [-12 – -18] | -15 [-11 – -18] | 0.584 |
| TR Vmax, m/s (n=57) | 2.5 [2.3 – 2.7] | 2.6 [2.4 – 2.7] | 0.132 |

Data are presented as median [interquartile range].
Abbreviations: A4Ch – Apical 4 Chamber View, GLS – Global Longitudinal Strain, LAVI –
Left atrial volume index, LVM index – LV mass indexed to Body Surface Area, MAPSE –
Mitral annular plane systolic excursion, PLAX – Parasternal long axis, RVEDD – Right
ventricular end diastolic diameter, RWT PW2 – Relative wall thickness posterior wall x 2;
TR Vmax – Maximum tricuspid valve regurgitation velocity. \* denotes <0.05, \*\* denotes

### 3.3 Myocardial mechanics

All PDF parameters are displayed in **Table 4** and **Figure 1**. Notably, both stiffness (*p* <0.001) and damping (*p*=0.012) increased at follow-up. However, there was no difference in load (*p*=0.97). Changes consistent with worsened mechanics of diastolic function could also be observed in E-wave maximal velocity, peak driving force, and peak resistive force (*p*<0.001 for all).

**Figure 1.**
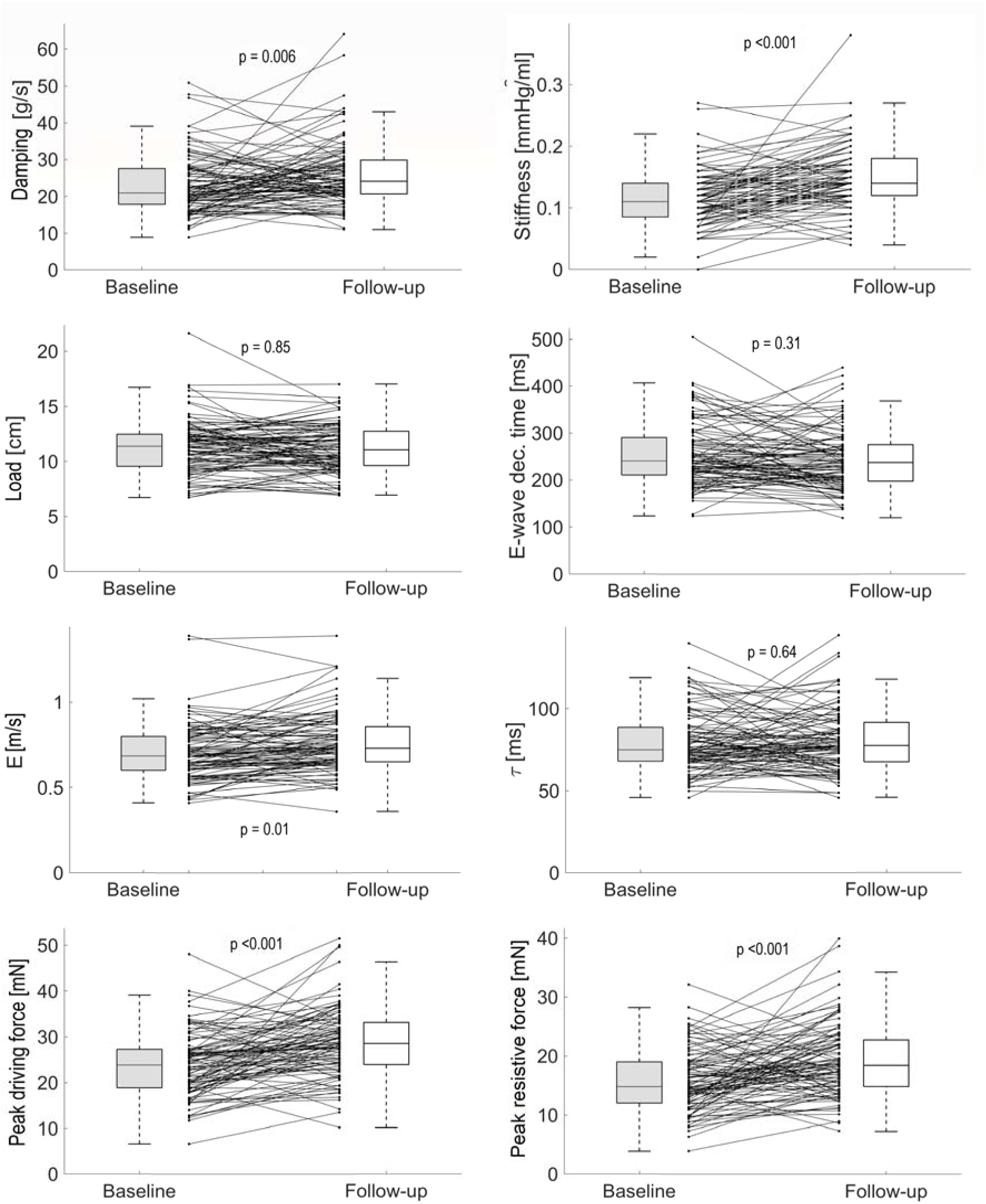
Left ventricular mechanics at baseline and follow-up. All PDF parameters at baseline (grey) and follow-up (white). Abbreviations: τ = Tau

**Table 4.** Change in PDF parameters between baseline and follow-up.

|  | Baseline (n = 96) | Follow-up (n = 96) | p-value | Normal reference<sup>#</sup> |
| --- | --- | --- | --- | --- |
| Stiffness, mmHg/ml | 0.11 [0.09 – 0.14] | 0.14 [0.12 – 0.18] | <**0.001** *** | 0.04 – 0.17 |
| Damping, g/s | 21.0 [18.0 – 21.0] | 24.2 [20.7 – 29.9] | **0.012** * | 10.2 – 25.3 |
| Load, cm | 11.4 [9.6 – 12.4] | 11.0 [9.6 – 12.7] | 0.970 | 6.3 – 14.4 |
| Peak driving force, mN | 23.8 [18.9 – 27.2] | 28.6 [24.0 – 33.2] | <**0.001** *** | 11.5 – 31.7 |
| Peak resistive force, mN | 14.9 [12.1 – 19.0] | 18.5 [14.9 – 22.8] | <**0.001** *** | 6.2 – 20.4 |
| Time constant of isovolumic pressure decay (tau), ms | 75 [68 – 88] | 78 [68 – 91] | 0.864 | 48 – 91 |
| Load independent index of diastolic filling (M), unitless | 1.11 [1.02 – 1.23] | 1.10 [1.00 – 1.24] | 0.791 |  |
| Intercept (B), mN | 5.7 [3.6 – 7.6] | 7.3 [4.5 – 9.5] | **0.010** * |  |

\* denotes <0.05, \*\* denotes <0.01, \*\*\* <0.001. <sup>#</sup> Normal reference values from the literature<sup>24</sup>.

No correlations with a correlation coefficient exceeding ±0.7 were observed between any PDF parameters and conventional echocardiographic variables, suggesting that conventional echocardiographic measures are not strong surrogates for the mechanistic measures quantified by the PDF method (see **Supplemental Table 1)**.

At baseline 58 patients (60 %) were characterized as having Normal phenotype, 24 (25%) as pEF, and 14 (15%) as rEF. In post hoc-analyses, PDF analysis revealed higher stiffness in the Normal and rEF-group and higher damping in all phenotype-groups. No difference in Load could be observed in any phenotype group (**Figure 2** and **Table 5)**.

**Figure 2.**
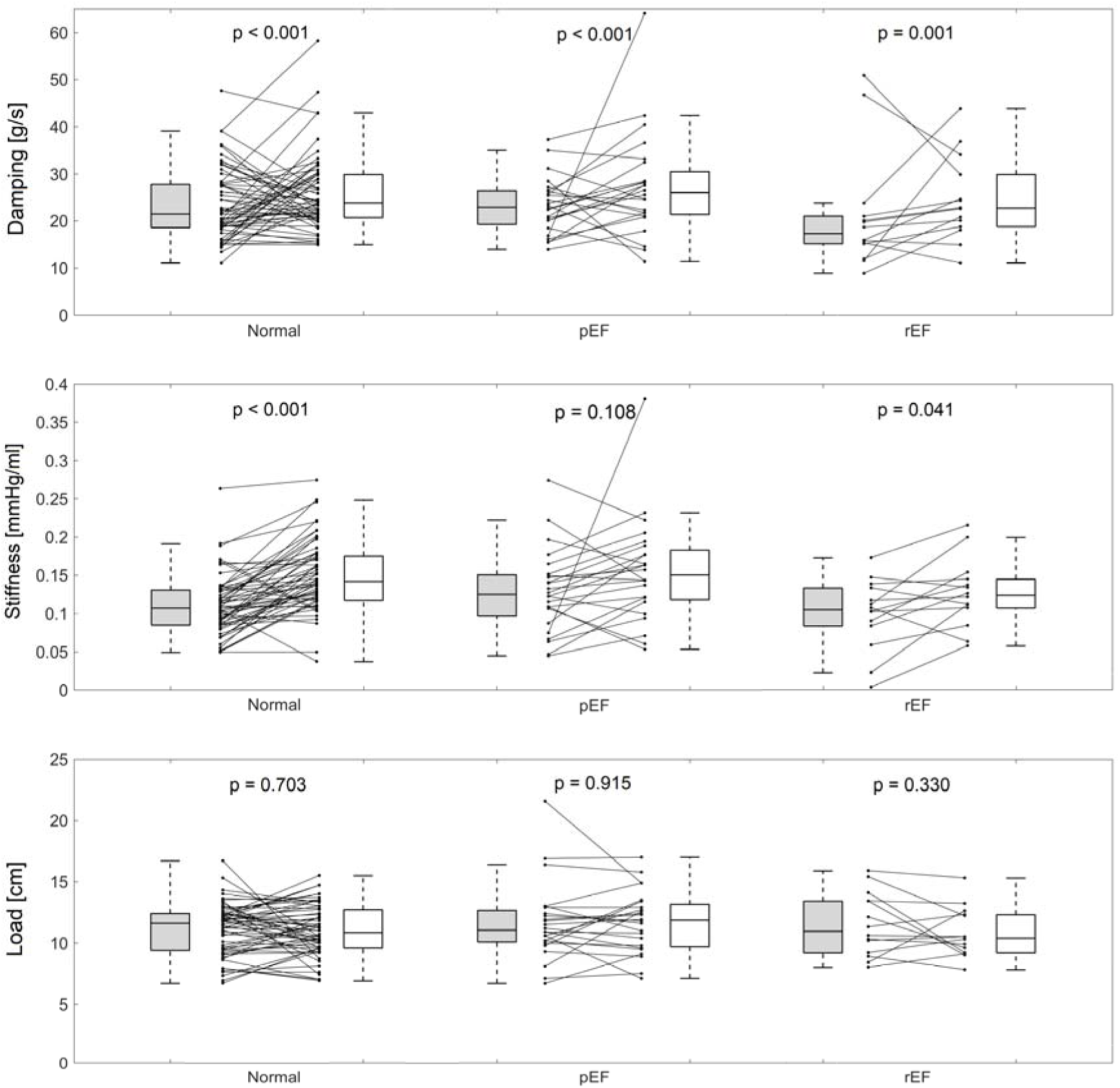
Left ventricular mechanics by cardiac phenotypes. The main PDF parameters divided by cardiac phenotypes at baseline (grey) and follow-up (white).

**Table 5.** Main PDF- and echocardiographic parameters at baseline presented by cardiac phenotype.

|  | **Normal (n = 58)** | **pEF (n = 24)** | **rEF (n = 14)** | **p-value for overall trend** |
| --- | --- | --- | --- | --- |
| Stiffness, mmHg/ml | 0.11 [0.08 – 0.13] | 0.13 [0.11 – 0.15] | 0.11 [0.08 – 0.13] | 0.178 |
| Damping, g/s | 21.5 [18.6 – 27.9] | 22.9 [18.9 – 26.5] # | 17.4 [14.4 – 21.7] # | 0.116 |
| Load, cm | 11.6 [9.4 – 12.4] | 11.1 [10.1 – 12.8] | 11.0 [9.1 – 13.6] | 0.937 |
| LVEDV, ml | 112 [97 – 127] § ‡ | 142 [113 – 155] § # | 165 [143 – 208] ‡ # | <0.001 *** |
| LVEDVi, ml/m<sup>2</sup> | 57 [48 – 66] § ‡ | 68 [59 – 77] § # | 85 [67 – 101] ‡ # | <0.001 *** |
| LVEF, % | 59 [57 – 63] ‡ | 61 [56 – 64] # | 40 [37 – 44] ‡ # | <0.001 *** |
| LV GLS, % | -19.0 [-20.0 – -17.7] ‡ | -17.2 [-20.4 – -15.8] # | -12.0 [-13.2 – -10.6] ‡ # | <0.001 *** |
| E/e', unitless | 7.8 [7.1 – 9.3] § ‡ | 10.7 [8.9 – 12.2] § | 12.0 [9.2 – 15.8] ‡ | <0.001 *** |
| LAVI, ml/m<sup>2</sup> | 32 [28 – 33] § ‡ | 41 [37 – 52] § | 37.2 [35.6 – 51.5] ‡ | <0.001 *** |
| NT-proBNP, ng/l<br>(n=91) | 148 [73 - 227] § ‡ | 273 [161 – 950] § # | 768 [466 – 1113] ‡ # | <0.001 *** |

### 3.4 Relationship between surgical procedure and postoperative LV function

No correlations with a correlation coefficient exceeding ±0.7 were observed between derived echocardiographic or mechanical parameters and the duration of cardiopulmonary bypass, aortic cross clamp time, or preoperative Euroscore2 (**Supplemental table 2)**.

## 4. Discussion

The main finding of this study is that at one year after elective revascularization by CABG, echocardiographic PDF analysis shows an overall mechanical stiffening and increased mechanical damping of the left ventricle, both indicating deterioration in the mechanics of diastolic function. Regarding damping, significant changes were seen in all cardiac phenotype groups while stiffness increased in the Normal and rEF-groups. The changes in conventional diastolic parameters were somewhat inconsistent and of small magnitude, and with no clear change in clinical assessment of diastolic function.

### 4.1 Diastolic left ventricular function following CABG

A number of studies have evaluated LV stiffness and associated biomechanical properties after surgical intervention. Notably, LV stiffness has been shown to increase immediately after CABG when quantified as the change in intraoperative end-diastolic pressure-area^25^ measured as the change in pulmonary artery wedge pressure per unit LV end-diastolic area measured by transoesophageal echocardiography. The acute impairment of diastolic function has been corroborated by other studies^26, 27^, but has been challenged by other work showing *decreased* regional stiffness in myocardium revascularized by CABG when defined as the regional work of the ventricle per unit volume of the revascularized myocardium three to four weeks after CABG^28^. In comparison, our study focused on assessing mechanical properties before and one-year post-CABG, seeking to define persistent changes manifesting relatively long-term post-intervention.

The current study showed no change in septal e’ and E/e’, and a low magnitude increase in lateral e’ one year after CABG. Furthermore, there was a concomitant increase in left atrial volume and reduction in left atrial strain. Taken together, the changes in these echocardiographic measures point towards a worsening of diastolic function. Previous studies using conventional echocardiographic variables reported an improved diastolic function within the first days after CABG but a return to preoperative values 18 months after surgery^29^. This considered, neither that study nor the current study were able to identify an improvement in diastolic function at one year when using conventional echocardiographic measures. The differences between this previous study and the current study likely arise from heterogeneities in acute post-operative response amongst patients with prior myocardial infarction, along with methodological differences in the assessment of diastolic function. Furthermore, in contrast to prior studies, the current study highlights persistent changes in myocardial stiffness one year after CABG. To the best of our knowledge, our study is the first to evaluate changes in LV diastolic function at the one-year postoperative timepoint in patients undergoing elective CABG. These results indicate persistent and continued deterioration in LV diastolic performance after CABG.

### 4.2 Differences between PDF and conventional diastolic parameters

The PDF method was chosen for analysis of diastolic function because of its ability to mechanistically describe LV relaxation. Moreover, tools for quantifying the PDF measures have improved following developments enabling rapid and precise semiautomatic measurement with excellent intra- and inter-reader reproducibility^7^.

The unique approach to diastolic LV functional assessment provided by the PDF method is the quantification of stiffness, damping, and load. Importantly, the differences observed in PDF parameters in our study were observed in the *absence* of changes in conventional echocardiographic measures of diastolic LV function including E/A, E/é, and E-wave deceleration time. This emphasizes the added mechanistic information provided by the PDF method, with early changes in mechanical behaviour possibly appearing prior to any obvious functional deterioration^10^.

Previous studies have been somewhat contradictory with regards to if, and to what extent, other diseases or conditions affect the main PDF parameters. Whilst the measures have been shown to be fairly uninfluenced by demographic parameters such as age and sex^10, 15^, conditions such as diabetes mellitus have been shown to more so increase stiffness and to a lesser extent damping, while hypertension has been found to increase both damping and stiffness^10^. These differences might be due to the selection of subjects based on different definitions of hypertension. Notably, diabetes and hypertension have only been shown to have a weak association with conventional echocardiographic diastolic parameters when adjusted for age^26^.

### 4.3 Potential mechanisms behind diastolic dysfunction following CABG

The role of the pericardium in cardiac relaxation is not fully understood^31^. The opening of the pericardium during CABG could provide a possible explanation for persisting post-operative alterations in LV relaxation, and further studies are warranted to clarify this mechanism. Another possible mechanism of action is that myocardial ischemia might not be the predominant cause of diastolic dysfunction, and as such, revascularization of the LV may not restore diastolic function^1^. This could be related to residual coronary microvascular dysfunction, or a combination of progression over time of the deleterious effects related to the risk factors associated with the need for revascularization. Especially among patients with preserved ejection fraction, diastolic dysfunction may develop as a result of diffuse interstitial myocardial fibrosis related to changes in the titin protein^32^. Taken together, the cause of postoperative deterioration in diastolic function and its clinical implications remains to be determined.

### 4.4 Limitations

The study cohort consisted of 90% male patients which was representative of the percentage of males undergoing isolated, on-pump CABG in Sweden during 2022, which was 84 %^34^. Sex differences regarding the changes in diastolic function following CABG could therefore not be assessed, and further future such investigations are justified. Since designing the study, new classification of heart failure have been published^35^. The definition of cardiac phenotype used in our study was based on well-established echocardiographic criteria and NT-proBNP and is compliant with the universal American College of Cardiology and American Heart Association definition of stages of heart failure^35, 36^. As assessed by the 2022 guidelines^35^, all patients in preoperative cardiac phenotype group Normal are considered to be in Stage A (“At-Risk for Heart Failure”) and all in the groups pEF and rEF in stage B (“Pre-Heart Failure”). For consistency, the PDF analysis was performed by a single observer, and this could be seen as a potential limitation. However, previous work from our group has highlighted excellent intra- and inter-reader reproducibility of the PDF method^7^, providing added confidence to the results of the current study. While it is acknowledged that the PDF method has a known measurement variability that may preclude meaningful individual clinical differentiation between health and disease^7^, the current study illustrates the robust utility of the method for gleaning mechanistic insight into differences between groups of sufficient sample sizes. With the assessment of early diastolic function provided by the PDF method using the mitral E-wave, no considerations are made to the possible contribution of atrial contraction as assessed by the mitral A-wave.

### 4.5 Conclusion

The current study found a deterioration in the mechanistic properties of diastolic function one year after CABG, seen as an increase in mechanical stiffness and damping while there were no or negligible changes in conventional echocardiographic measures of diastolic LV dysfunction. This work highlights the complexity of cardiac diastolic remodelling, and the value of including mechanistic evaluation by methods beyond conventional echocardiographic measures to understand changes in mechanical LV relaxation.

### 4.6 Clinical perspective

It is unclear whether or not CABG has beneficial effects on diastolic dysfunction. Conventional echocardiographic measures of diastolic function cannot characterize the mechanics of diastolic function. The use of a mechanistic approach to analysing LV relaxation using the echocardiographic PDF method was able to identify consistent deterioration in diastolic function one year after CABG in a way that is not clinically apparent by conventional echocardiographic evaluation.

## Data Availability

All data produced in the present study are available upon reasonable request to the authors.

## Data availability

The data underlying this article will be shared on reasonable request to the corresponding author.

## 8. Supplemental materials

Supplemental tables S1 – S2

