## Supplemental tables 1 and 2 for "Deteriorated mechanics of left ventricular diastolic filling one year after coronary artery bypass grafting"

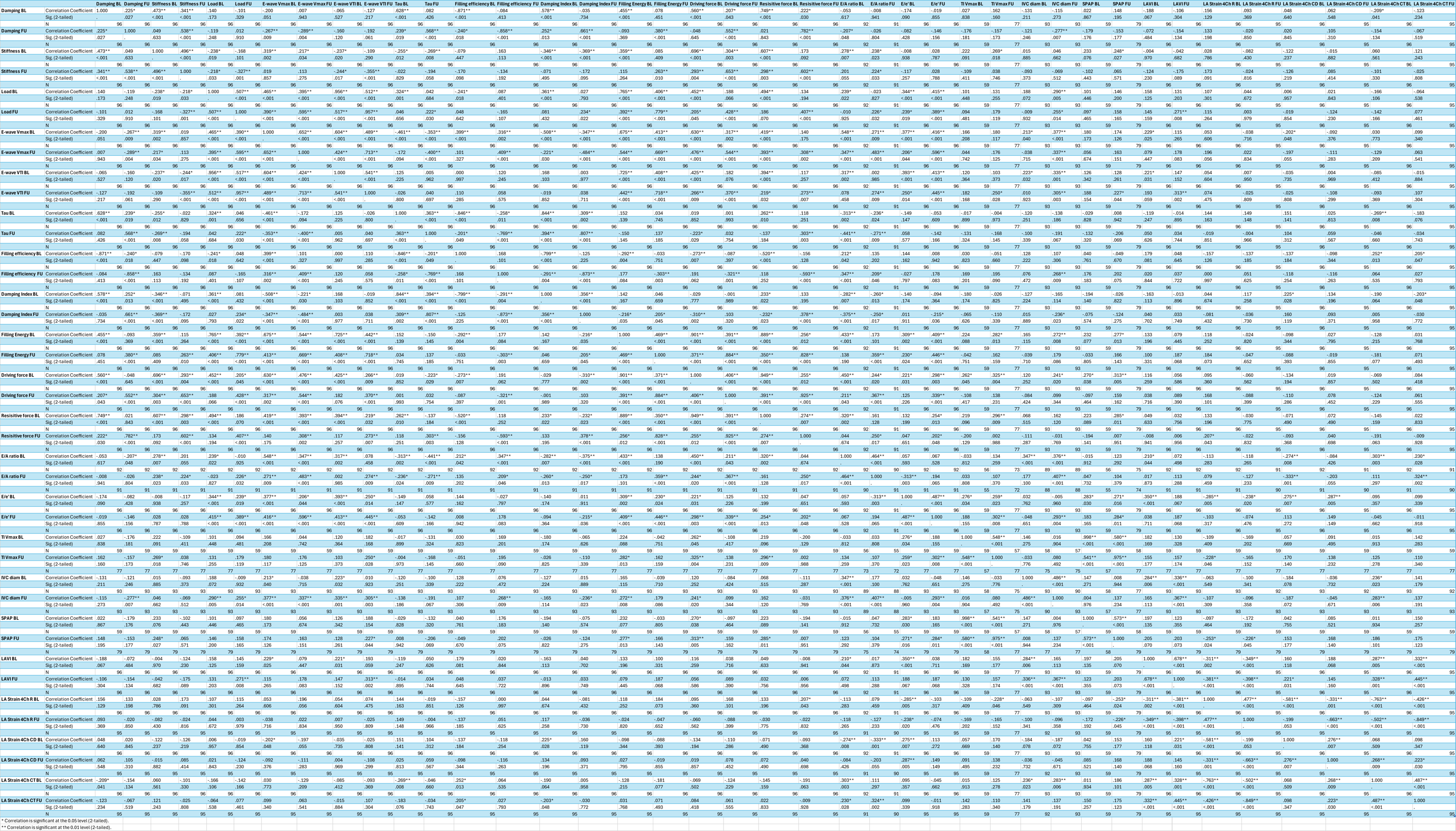

**Supplemental Table 1**. – Correlations between PDF parameters and conventional diastolic echocardiographic parameters.

**Supplemental table 1**

Presented as Spearman’s correlation. Abbreviations: BL – baseline, FU – follow-up, Sig (2-tailed) – two tailed statistical significance, * – alpha level of 0.05, ** – alpha level of 0.01, N – number of patients, VTI – velocity time integral (cm), E/A-ratio – ratio between early and atrial wave of mitral inflow (measured using pulsed wave (PW) Doppler), E/e’ – ratio between early transmitral flow (measured using PW Doppler) and mean of septal and lateral e’ (measured using tissue velocity imaging by PW Doppler), TI Vmax – maximal velocity (m/s) of tricuspid regurgitation, IVC diam – diameter of inferior vena cava (mm), SPAP – estimated systolic pulmonary artery pressure (mmHg), LAVI – left atrial volume indexated to Body Surface Area (ml/m^2^), LA Strain 4Ch R – left atrial reservoir strain in apical four chamber view (%), LA Strain 4Ch CD – left atrial conduit strain in apical four chamber view, LA Strain 4Ch CT – left atrial contraction strain in apical four chamber view.

**Supplemental Table 2**. Correlations between change in PDF parameters, conventional echocardiographic parameters and perioperative surgical parameters.

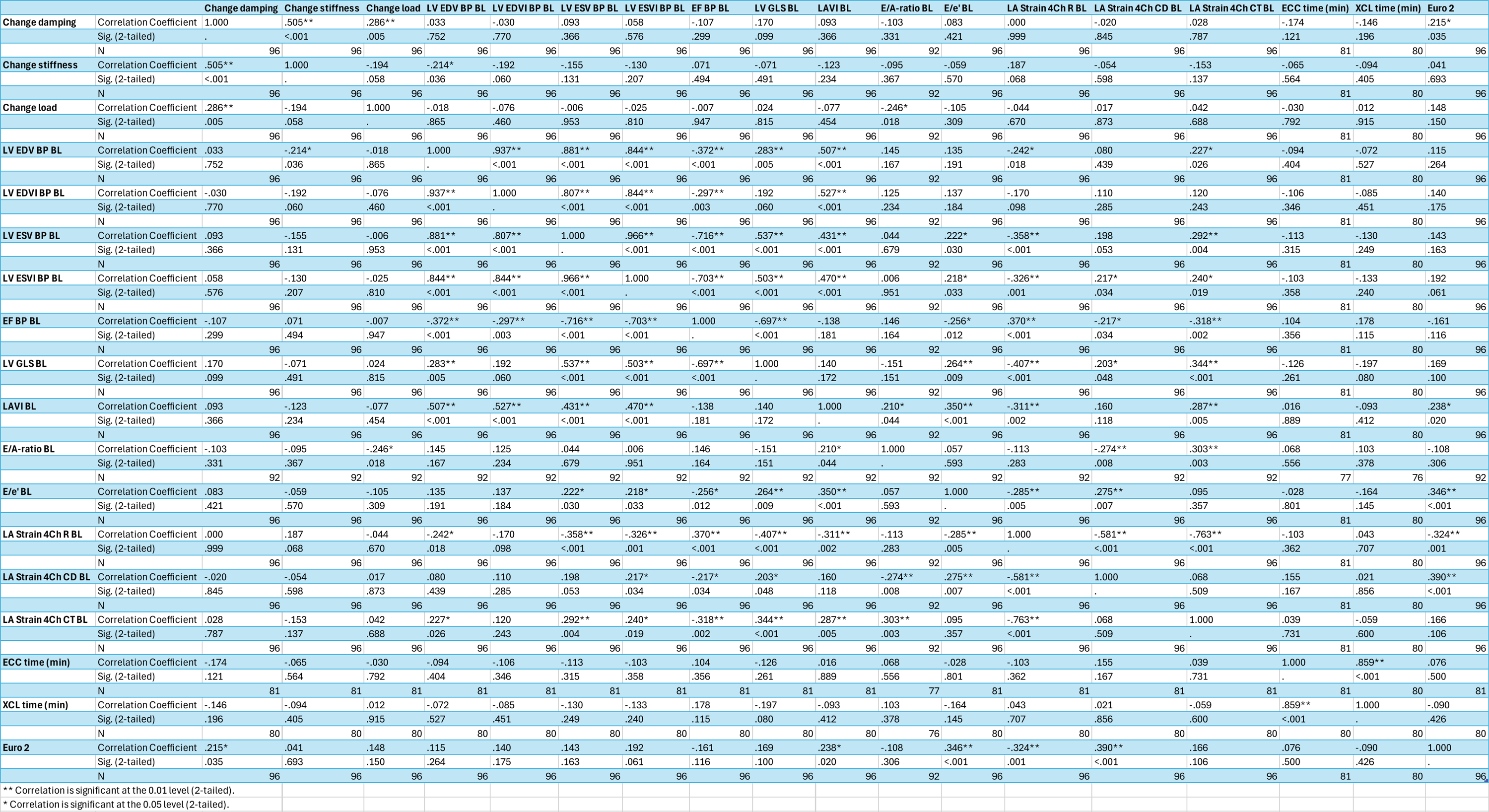

**Supplemental table 2**

Presented as Spearman’s correlation. Abbreviations: BL – baseline, FU – follow-up, Sig (2-tailed) – two tailed statistical significance, * – alpha level of 0.05, ** – alpha level of 0.01, N – number of patients, Change – change in parameter between BL and FU (calculated as FU-BL), LV EDV BP – left ventricular (LV) end diastolic volume (calculated using Simpson Biplane), LV EDVI – LVEDV BP indexated to body surface area (BSA), LV ESV BP – LV end systolic volume (calculated using Simpson Biplane), LV ESVI BP – LV ESV BP indexated to BSA, EF BP – LV ejection fraction (calculated using Simpson Biplane), LV GLS – LV Global Longitudinal Strain (calculated from apical 4-, 2- and 3-chamber views), E/A-ratio – ration between early and atrial wave of mitral inflow (measured using pulsed wave (PW) Doppler), E/e’ – ratio between early transmitral flow (measured using PW Doppler) and mean of septal and lateral e’ (measured using tissue velocity imaging by PW Doppler), LA Strain 4Ch R – left atrial reservoir strain in apical four chamber view, LA Strain 4Ch CD – left atrial conduit strain in apical four chamber view, LA Strain 4Ch CT – left atrial contraction strain in apical four chamber view, ECC time – duration of extracorporeal circulation, XCL time – duration of aortic cross clamping, Euro 2 – preoperative Euro2-score.
